# Introducing causal inference to the medical curriculum using temporal logic to draw directed acyclic graphs

**DOI:** 10.1101/2020.08.02.20166900

**Authors:** George T.H. Ellison

**Affiliations:** Centre for Data Innovation, Faculty of Science and Technology, University of Central Lancashire, Preston, PR1 2HE UK

**Keywords:** Directed acyclic graph, DAG, causal inference, statistical modelling, observational data, temporality, MBChB, undergraduate

## Abstract

Directed acyclic graphs (DAGs) might yet transform the statistical modelling of observational data for causal inference. This is because they offer a principled approach to analytical design that draws on existing contextual, empirical and theoretical knowledge, but ultimately relies on temporality alone to objectively specify probabilistic causal relationships amongst measured (and unmeasured) covariates, and the associated exposure and outcome variables. While a working knowledge of phenomenology, critical realism and epistemology seem likely to be useful for mastering the application of DAGs, drawing a DAG appears to require limited technical expertise and might therefore be accessible to even inexperienced and novice analysts. The present study evaluated the inclusion of a novel four-task directed learning exercise for medical undergraduates, which culminated in temporality-driven covariate classification, followed by DAG specification itself. The exercise achieved high levels of student engagement, although the proportion of students completing each of the exercise’s four key tasks declined from close to 100% in tasks 1 and 2 (exposure and outcome specification; and covariate selection) to 83.5% and 77.6% in the third and fourth tasks, respectively. Fewer than 15% of the students successfully classified all of their covariates (as confounders, mediators or competing exposures) using temporality-driven classification, but this improved to more than 35% following DAG specification – an unexpected result given that all of the DAGs displayed at least one substantive technical error. These findings suggest that drawing a DAG, in and of itself, increases the utility of temporality-driven covariate classification for causal inference analysis; although further research is required to better understand: why even poorly specified DAGs might reduce covariate misclassification; how ‘wrong but useful’ DAGs might be identified; and how these marginal benefits might be enhanced with or without improvements in DAG specification.

## “It’s such a mistake, I always feel, to put one’s trust in technique”

**(George Smiley in: *The Looking Glass War* by John Le Carre, 1965)**

## Introduction

### Statistical skills at the heart of evidence-informed policy and practice

Notwithstanding George Smiley’s caution and the fallibility of method, statistical expertise plays an increasingly important role in generating and interpreting quantitative evidence to inform policy and practice (Teater et al. 2017; Flyberg et al. 2019); and training in statistical skills has always needed to keep pace with ongoing developments in analytical practice (Tu and Greenwood 2012; Porta et al. 2015; Efron and Hastie 2016; Hokimoto 2017). Indeed, perhaps the most important contribution that statistics can make to evidence-based decision-making (and, by extension, the contribution that statistical skills training can make to professionals and the lay public, alike) is in revealing and dealing with the many different sources of bias that can occur when analysing and interpreting obvious, observable differences between ostensibly comparable phenomena (Flyberg et al. 2019). Regardless of whether such comparisons are an integral part of human nature or a conditioned response to our natural and social environments (Ross 2019), they manifest as compelling objects of enquiry and speculation even for those trained to recognise how method, context and perspective often determine the extent to which these comparisons actually provide precision, generalisation and causal/predictive insight, respectively (Asprem 2016).

In the not so recent past, concern with selection bias (and, to some extent, with chance associations generated by underpowered sample sizes) led to a renewed focus on statistical training in: the use of research design (particularly experimentation and randomisation) to address potential sources of bias when estimating evidence of cause and effect; and on critical appraisal and research synthesis techniques (including meta-analysis) to deal with contradictory findings from different studies undertaken in different contexts using different techniques (Djulbegovic and Guyatt 2017). Over time, the limited utility of this approach has led to a resurgence of interest in the analysis of non-experimental (observational) data and the synthesis of ‘real world evidence’ (e.g. Klonoff 2020; though see also: Losilla et al. 2018). Indeed, there is broad consensus that such analyses will remain far more common in many applied disciplines, not least in the era of ‘medical tech’ and ‘Big Data’ (Eustace 2018); and in contexts where intervention complexity, ethics, governance, safety, participation and cost make experimentation impossible or simply undesirable and undesired (Meyer et al. 2019).

### Why observational data, why causal inference and why directed acyclic graphs?

Many of the same sources of bias that led to the earlier focus on experimentation and randomisation – particularly those relating to confounding and sampling bias – continue to threaten the validity of observational analyses, not least because design constraints mean such studies exert little control over the allocation of naturally occurring ‘exposures’ (be these physical, biological or social phenomena). Nonetheless, recent efforts to address confounding and sampling bias have made substantial progress; and the emergence of ‘causal inference’ as a novel interdisciplinary field spanning statistics, mathematics and computing (as well as the applied social and biomedical sciences), has helped in the translation of abstract theoretical techniques into accessible and practical applications (Porta et al. 2015).

One technique at the centre of these developments, which can help to guide and reveal the otherwise implicit nonparametric considerations that underpin the analysis of observational data, is the ‘directed acyclic graph’ or DAG. This is a form of causal path diagram that seeks to represent what the analyst believes to be the underlying ‘data generating mechanism’ – a somewhat abstract and quasi-critical realist concept (Spanos 2010) that nonetheless encapsulates how mathematical and statistical analyses of the joint information provided by data from each of the measured variables (the so-called ‘known knowns’) might offer insights into the processes responsible for creating these (the ‘known unknowns’ and potential ‘unknown unknowns’ (Recker 2015). In practice, the data generating mechanism is rarely (if ever) understood with absolute certainty, not least for data sets in which complex social processes are at play (whether in the variables and contexts concerned, or simply in the cultural practices involved in their scientific conceptualisation, operationalisation and metricisation). Yet while the speculative theoretical nature of DAGs may not necessarily reflect reality, their use helps to make explicit a critical step that is often only implicit within established analytical practices (Law et al. 2012), and makes it possible to separate out: the theory being tested; and the statistical analyses required to evaluate this.

DAGs therefore offer an immediate and compelling contribution to improving the analysis of observational data because they aim to facilitate the elucidation of plausible (yet imperceptible) data generating processes and reveal many of its critical features so that these can inform the design of statistical models and ensure these are capable of providing robust effort for causal inference. In the process, DAGs enable analysts to summarise and share *both*: their theoretical understanding, beliefs and speculation regarding the true nature of the underlying data generating mechanism; *and* how this is reflected in the design of the statistical analyses intended to evaluate this. In effect, using DAGs to strengthen the design of such analyses requires analysts to up their game by thinking through their assumptions and taking greater care to prepare, double check and open these up for scrutiny, debate and challenge (Textor et al. 2016).

These advantages are matched by the ease with which DAGs can help analysts seeking causal inference to identify which of their variables play important roles in the ‘focal relationship’ (or ‘relationship of interest’ (Tennant et al. 2017): i.e. the extent to which a hypothesised cause (the ‘specified exposure’) might genuinely affect a particular consequence (the ‘specified outcome’). Indeed, beyond the variables specified as the ‘exposure’ and ‘outcome’ in any given analysis, each of the other measured (and all of any other *unmeasured* or *latent*) variables can act as: ‘confounders’, ‘mediators’, ‘competing exposures’ or ‘consequences of the outcome’ (see Figure 1). Confounders are covariates that cause *both* the specified exposure and the specified outcome and which, in the absence of adjustment, can reverse, enhance or mask the direction, strength and precision of the true relationship between exposure and outcome (as a result of ‘confounder bias’ (VanderWeele 2019). Mediators are covariates that are caused by the specified exposure and cause the specified outcome (i.e. they fall along one potential causal path between the specified exposure and outcome). Like confounders, mediators can reverse, enhance or mask the direction, strength and precision of the true relationship between exposure and outcome (as a result of inferential bias known as ‘mediator bias’ (Richiardi et al. 2013) – though, unlike confounders, mediator bias only occurs *after* mediator adjustment. Meanwhile, competing exposures are covariates that are causally unrelated to the specified exposure but which cause (and can therefore explain a proportion of the variance in) the specified outcome. Adjustment for genuine competing exposures has no effect on the strength of the relationship observed between the specified exposure and outcome, but can improve the precision of this relationship (Tennant et al. 2017). Finally, consequences of the (specified) outcome are covariates that do not necessarily have any causal relationship with the specified exposure but are caused by the specified outcome. Like mediators, adjustment for these variables can reverse, enhance or mask the direction, strength and precision of the true relationship between exposure and outcome as a result of biases that are essentially the same as those generated through ‘conditioning on the outcome’

**Figure 1.**
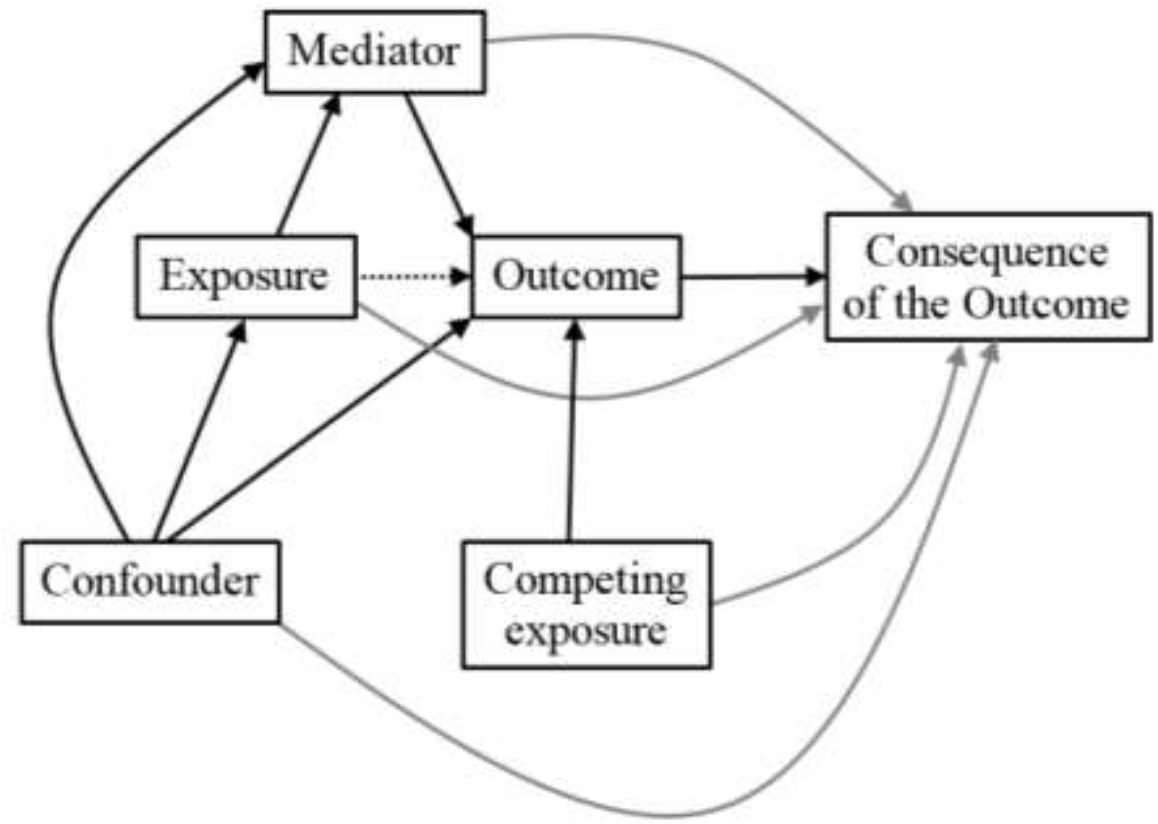
A stylised example of a directed acyclic graph (DAG) illustrating: (i) the four principal roles that covariates (represented as ‘nodes’) can play, namely: confounders; mediators; competing exposures; and consequences of the outcome; and (ii) the unidirectional probabilistic causal paths (represented as ‘arcs’ with arrows) between consequential nodes and all causal node(s) that temporally precede them.

Clearly, by helping analysts better understand which covariates warrant adjustment in statistical models examining the direction and strength of potential causal relationships amongst measured variables in observational datasets, DAGs can not only transform their own analytical practices but can also enhance their ability to critique and learn from the theories and modelling practices of others. These improvements in analytical modelling to support causal inference from observational data have helped to transform what passed for accepted/acceptable practice where, until relatively recently: there was even little consensus on how to define or identify a true confounder (VanderWeele and Shpitser 2013); arbitrary and ostensibly haphazard techniques (including those based simply on the covariates for which data were available (Schelchter and Forsythe 1985) were commonplace; and even the more reputable parametric techniques were deeply flawed, such as:

i. selecting covariates for adjustment on the basis that they display strong univariate correlations with *either* the specified exposure *and/or* the specified outcome; and
ii. using step-wise techniques to select a group of covariates whose adjustment optimises the total amount of variance explained by the model.

Indeed, since covariates acting as confounders, mediators, competing exposures and consequences of the outcome can display *both* strong *and* weak univariate correlations with specified exposures and outcomes (i); and since adjustment for each type of covariate can both strengthen and weaken the total amount of variance explained by the model (ii), neither of these parametric techniques are capable of distinguishing which covariates might act as *genuine* confounders, and which are likely to act as mediators, competing exposures or consequences of the outcome. DAGs have addressed this impasse by providing a principled schema based on two key tenets of temporal logic, namely that: any cause must precede its consequences (hence ‘directed’); and no consequence can influence any of its own cause(s) (hence ‘acyclic’).

DAGs greatly improve the ability of analysts to identify potential confounders and include these in the ‘covariate adjustment sets’ required to mitigate the effect of confounder bias in observational analyses which aim to support causal inference. DAGs have also provided the tools required to identify, better understand and explicate hitherto complex and challenging sources of bias in observational bias – perhaps the most famous of which is the ‘low birth weight paradox’ (the observation that low birth weight babies are more likely to survive if their mothers had smoked during pregnancy), which DAGs have revealed to be simply an example of ‘selection collider bias’ (Porta et al. 2015). Nonetheless, the utility of DAGs to improve the design of observational analyses for causal inference ultimately depends upon the analyst’s contextual understanding and the availability of measured covariates, and this will often require careful thought to correctly identify which of these covariates occur before the specified exposure (and can therefore be considered potential confounders as probabilistic causes of *both* the exposure *and* the outcome). These considerations aside, drawing a DAG appears deceptively simple and seems to require limited technical expertise. As such, might not DAG specification and its tangible benefits be accessible to even inexperienced and novice analysts?

This then was the rationale behind including DAGs in the statistical training provided to undergraduate medical students at the University of Leeds (Ellison et al. 2014a), which since 2012 has involved lectures explaining the theory behind DAGs and the rationale for using these in the analysis of observational data to support causal inference. It has subsequently included directed learning exercises to provide practical instruction in the DAG specification using a range of different techniques (including: graphical, cross-tabulation and relational; Ellison et al. 2014b) to critically appraise the covariate adjustment set used in a previous flawed analysis published by the author and former colleagues (Harris et al. 1999). Evaluations of this approach, and the different techniques involved, confirmed that the vast majority of students found the concept of DAGs intuitive, and soon learned to correctly identify potential confounders, likely mediators and competing exposures (and which of these to include, and exclude, in the covariate adjustment sets required to minimise the impact of bias from available/measured confounders; Ellison et al. 2014a). However, many students also found the use of a published example (on a topic about which they knew little) limited their contextual understanding; while others found the traditional (graphical) DAG specification technique challenging and time-consuming (particularly when large number of covariates were involved, and the DAGs concerned became cluttered with nodes and arcs). Efforts to simplify DAG specification using cross-tabulation and relational approaches to systematically examine every possible relationship between each pair of covariates appeared to have little impact on the difficulty and time involved; and rather than enabling students to focus on temporality as the basis on which to objectively and systematically assign likely (probabilistic) causal relationships between one covariate and another, many students fell back on their prior knowledge, experience, training and beliefs to postulate which of the possible relationships between measured covariates and the specified exposure and outcome were likely to be causal.

To address concerns regarding contextual understanding, complexity and the amount of time required to complete the directed learning exercise, in subsequent years DAG specification was incorporated into the course’s Project Protocol assignment. This enabled students to choose the context, topic and focus of a hypothetical clinical project designed to assess the potential causal relationship between a modifiable aspect of health care delivery (the exposure) and compliance with an established best practice guideline (the outcome). At the same time, to reduce the directed learning exercise’s complexity (and the amount of time required to complete this), students were directed to select just eight “potentially important variables… that are likely to cause the[ir selected] outcome” for subsequent consideration as potential confounders, likely mediators and competing exposures – far fewer than the 28 covariates included in the previous cross-tabulation and relational approaches used. Finally, an additional task was introduced into the directed learning exercise to ensure that students drew on temporality-related considerations to classify covariates as potential confounders, likely mediators or competing exposures *before* they attempting to specify their DAG in graphical form. The aim of the present study is therefore to evaluate the impact of these changes on: student engagement and task completion; the (mis)classification of covariates; the (mis)specification of DAGs; and the likely utility of each task for improving the design of observational analyses to support causal inference.

## Methods

### The Research, Evaluation and Special Studies Strand at Leeds Medical School

Leeds Medical School introduced its novel *Research, Evaluation and Special Studies* (RESS) strand in 2010 in response to the 2009 update of *Tomorrow’s Doctors* by the UK’s General Medical Council (GMC 2009), which included a focus on “The Doctor as a scholar and a scientist” as the first of three overarching outcomes. The School’s response involved a radical refresh of the undergraduate curriculum (Roberts 2011), including a focus on involving undergraduate medical students in research (Murdoch-Eaton et al. 2010). RESS sought to deliver this vision through a spiral strand spanning years 1 through 5 involving successive modules in years 1, 2 and 3, and culminating in an 18-month *Extended Student-selected Research & Evaluation Project* (ESREP) in years 4 and 5.Core research-relevant principles, practices and responsibilities are introduced in the first year RESS1 module (including hands-on research project experience), whilst the second year RESS2 module focuses on core statistical and analytical skills and their application to open-source population health data. These skills are synthesised, consolidated and extended in the third year RESS3 module, with a particular focus on sources of bias in applied clinical and health services research, and on accessible techniques for addressing these through: randomisation; sample size estimation; standardised data collection/extraction; and confounder adjustment using multivariable statistical analysis. The last of these is where a directed learning exercise focussing on DAG specification was first introduced into the MBChB curriculum in 2012 (Ellison et al., 2014a; 2014b).

In preparation for the Extended Student-selected Research & Evaluation Project (ESREP) in years 4 and 5, the final assignment for the RESS3 module was designed in the form of a detailed Project Protocol. This involves students designing a hypothetical service evaluation project to examine the relationship between ostensibly modifiable health service characteristics (such as: the context of health care delivery; the equipment and facilities available; and the experience, expertise and training of the health care professionals involved) on a specified health service/healthcare outcome. This design sought to equip all students with the skills required to evaluate the potential impact of changes in health service delivery on subsequent health (and healthcare) outcomes – skills considered core to all qualified clinicians (GMC 2009). As a result, this design also provided a level of consistency sufficient to permit the evaluation of skills development while allowing students substantial choice in the context, topic and focus of their chosen project; an important consideration given the impact of student engagement on high intensity skills acquisition (Miller et al. 2011).

### The RESS3 directed learning exercise on DAG specification

The RESS3 module begins with a series of lectures explaining the distinction between clinical audit and service evaluation, and between biomedical and methodological research (‘discovery’) and applied health service research (‘translation’). These sessions also offer practical guidance on: expert and stakeholder involvement; institutional governance; and ethical approval procedures; and seek to emphasise and reinforce how these preparatory steps can offer tangible benefits in helping to establish: what mistakes to avoid; which questions to prioritise; what support might be required; and any necessary constraints to protect researchers and research participants alike. These lectures conclude with peer-led training in the use of *NICE Evidence Search* (a database developed by the National Institute for Clinical and Care Excellence to facilitate access to selected authoritative evidence on health and social care, including best practice guidelines), delivered with support from the NICE Evidence Search Champions Scheme (Rowley et al. 2015; Sbaffi et al. 2015).

The students then focus their attention on developing their Project Protocol for a hypothetical audit-cum-service evaluation study that aims to support robust causal inference for improving adherence to a selected best practice guideline. The design of this assignment aims to balance the consistency required for the assessment of learning, with opportunities for students to choose, innovate, succeed and excel. To this end, the RESS3 Project Protocol assignment requires students to specify, as their project’s ‘specified outcome’, a clinical practice guideline in any specialty or context of interest to them. Likewise, for their ‘specified exposure’, students can then choose any aspect of health service organisation/delivery that might feasibly affect adherence to their selected practice guideline.

Over the weeks that follow subsequent lectures, large-group lectorials and small-group tutorials support the students to develop each of the five key skills required to design a detailed Project Protocol that is capable of generating the evidence required to support robust causal inference regarding the nature, direction and strength of any relationship between their specified exposure and specified outcome. These skills involve: selecting which additional variables (i.e. covariates) are likely to be (un)necessary to measure, collect or extract; developing standardised data measurement and/or extraction procedures to strengthen precision and internal validity; designing coherent inclusion and exclusion criteria to optimize both internal and external validity; conducting sample size estimation to generate a suitably powered sampling strategy; and selecting an appropriate covariate adjustment set to minimise bias from available/measured confounders and thereby inform the design of suitable multivariable statistical models to support causal inference and interpretation.

The directed learning exercise developed to support the acquisition of the last of these skills draws together four successive tasks which are summarised on a two-side worksheet that students work through during the lectorial following their introductory lecture on causal inference and DAGs; and subsequently discuss in supervised small-group tutorials later that same day. These four tasks comprise:

Task 1 – *Exposure and Outcome Specification:* In this task, students first choose an appropriate exposure variable which “aims to measure/record the variation in clinical practiced experienced by” service users receiving care within the clinical context(s) chosen by the student. Students then choose a suitable outcome variable which describes “whether each patient in your proposed study has received care that complied with the NICE guidance/standard” as chosen by the student.
Task 2 – *Covariate Selection:* This task involves the selection of what are described as “potentially important variables – excluding the ‘exposure’ – that are likely to cause the ‘outcome’”, for which students are reminded that “such causes *must* precede the variable they cause – in this instance they must precede the ‘outcome’ and cannot be a subsequent ‘consequence of the outcome’”
Task 3 – *Temporality-driven Covariate Classification:* Students are once more reminded that “‘causes’ *must* temporally precede the variable they cause” before they are directed to identify: which of the covariates selected during task 2 are confounders (since they were “ALSO likely to cause the ‘exposure’”); which are mediators (because they were “ALSO likely to BE CAUSED BY the ‘exposure’”); and which must be competing exposures (on the basis that they are neither causes of, nor are they caused by, the specified exposure).
Task 4 – *DAG Specification:* Finally, in the last of the four tasks the students are simply directed to “sketch a Directed Acyclic Graph that includes your… ‘exposure’, ‘outcome’ and each of the…” selected covariates as identified and classified during task 2 and 3 (above), respectively.

### Engagement, completion, covariate (mis)classification and DAG (mis)specification

To evaluate the directed learning exercise and each of its subsidiary tasks, lectorial worksheets were anonymised, photocopied and the original returned to the students concerned. The anonymised worksheets were then used to assess the proportion of students who had successfully completed each of the four tasks (as outlined above) as a primary indicator of student engagement. The medical (sub)specialities pertinent to the clinical contexts, topics and foci chosen by each student were then classified and enumerated to provide an assessment of the extent to which students had been able to exercise choice when selecting these. Similar summaries of the guideline-related outcomes, ‘modifiable’ health service exposures and speculative causes of each student-selected outcomes (i.e. the covariates selected in the second task) offered an assessment of the extent to which students explored different healthcare pathways and the factors that might influence these. Subsequent, in-depth assessment of the temporality-driven covariate classifications completed during the third task (and those reflected by the DAGs specified in the fourth task) permitted the estimation of covariate misclassification rates in each of these tasks, disaggregated by the type of covariate concerned (i.e. by confounder, mediator and competing exposure). Finally, each of the specified DAGs were subjected to detailed examination to calculate the average number of variables (nodes) and causal paths (arcs) these contained, and to enumerate the frequency of unorthodox features and technical errors (such as the use of ‘super-nodes’, the unwarranted omission of arcs, the use of undirected or bidirectional arcs, and the presence of cyclical paths). Together, these analyses aimed to evaluate the potential utility of the directed learning exercise for strengthening the analytical knowledge, skills and competencies of undergraduate medical students in the development of statistical models to support causal inference from observational data (such as routinely collected health service data on which many of their exposures, outcomes and covariates were likely to rely).

## Results and discussion

### Completion of the four successive tasks in the directed learning exercise

A total of 85 anonymised worksheets were available for analysis in the present study. Most of the students involved (58; 68.2%) had successfully completed all four of the tasks in the directed learning exercise, although completion rates declined with each successive task from: close to 100% for the first and second task (exposure and outcome specification, and covariate selection, respectively); to 83.5% (71/85) for the third task (temporality-driven covariate classification); and 77.6% (66/85) for the fourth and final task (DAG specification). Thus, all but one of the students (98.8%) identified both: a suitable outcome (relevant to an established clinical guideline; with/without an associated health service target); and an appropriate exposure (comprising a discrete component or characteristic of clinical care that temporally preceded, and was a plausibly modifiable cause of, their specified outcome). Indeed, the single student who failed to complete this task simply appeared to have mis-labelled their outcome and exposure, since their subsequent DAG correctly positioned these variables in their most likely temporal sequence (i.e. the listed exposure had become the outcome and vice versa).

All 85 of the students also went on to generate a list of “potentially important variables… that are likely to cause the[ir specified] outcome”; the median number of such covariates being 8 with a range of 5 to 14. However, only 71 of the students (83.5%) disaggregated these lists of covariates into those they considered potential confounders (median number: 3; range: 1-8), likely mediators (median number: 2; range: 0-6) and/or competing exposures (median number: 2; range: 0-6); and an even smaller proportion of students (68; 80.0%) completed a sketch of their DAG based on their specified exposure and outcome, and on the list covariates they felt likely to cause their selected outcome.

These findings suggest that, while most of the students were able to complete all four tasks, a growing proportion found the last two tasks (temporality-driven covariate classification, and DAG specification) more challenging and difficult to complete within the time available. Indeed, since a small proportion of the students who completed the fourth task (DAG specification: 8/66; 12.1%) did so *without* completing the third task (temporality-driven covariate classification), it seems likely that the latter was experienced as the most difficult, time-consuming and, perhaps, least important for completing the final (DAG specification) task. This last possibility is worth exploring further if, as seems plausible, the conceptual challenge involved when classifying covariates on the basis of their temporal relationship with the specified exposure (while ignoring any potentially erroneous prior knowledge or belief regarding their causal/functional relationships therewith), meant it was possible to complete the third task but not always the fourth as well; while, in contrast, the fourth task could be completed without completing the third. Further insight into this possibility is explored later in the analyses that follow (see below).

### Student-selected clinical contexts, outcomes, exposures and covariates

The practice guideline-related outcomes chosen by students during the first task (exposure and outcome specification) in the present study spanned a wide range of clinical specialties; and the only notable omissions were public health, oncology, radiology, intensive care, pathology, anaesthesia and surgery (see Table 1). The guidelines themselves likewise covered every stage of the healthcare pathway, the commonest being: assessment and diagnosis (17); referral, monitoring and follow-up (20); and the provision of advice, medication, therapy and/or care (30). Somewhat unsurprisingly, given these outcomes were derived from practice guidelines that commonly serve as performance criteria, most (58; 68.2%) had associated delivery/waiting time targets.

**Table 1.**
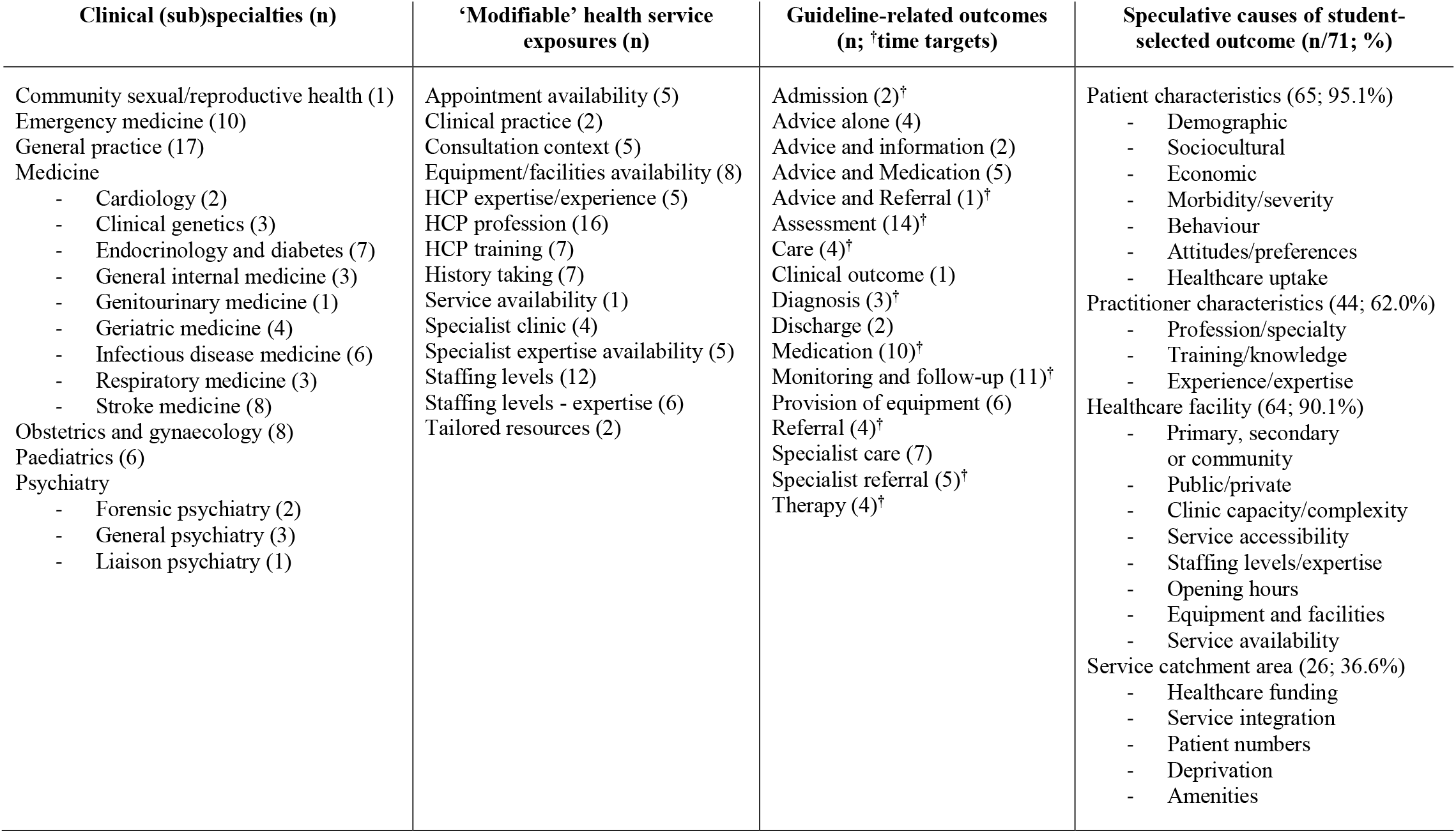
Clinical specialties, ‘modifiable’ health service exposures, and guideline-related outcomes selected by 85 third year MBChB students receiving instruction in the use of DAGs to inform analytical modelling for causal inference. Students were also invited to speculate what additional covariates might act as likely causes of their selected outcome (in addition to their specified exposure), and these have been classified under four headings: patient/practitioner characteristics; healthcare facilities; and catchment.

Meanwhile, the preceding health service characteristics considered amenable to ‘modification’ (and possible subsequent intervention) included: the *location* of health care delivery; the *staff* involved in delivering this care; and the *expertise* and *equipment* required/available. Of these, the most common involved consideration of which professions were available to/involved in the delivery of care (16); the training, experience and expertise of the health care practitioners involved (17); and associated staffing levels and staff-to-patient ratios (18). These features also predominate amongst the various patient-, practitioner-, facility- and catchment-specific parameters which students speculated might act as potential causes of their selected outcomes during the second, covariate selection task; although a far greater proportion of students included those covariates specific to patients (95.1%) or health care facilities (90.1%) than to either practitioners (62.0%) or health service catchments (36.6%; see Table 1).

Clearly, despite the tightly structured design of the directed learning exercise, students were able to exert substantial choice when selecting the clinical specialties and contexts in which to situate their hypothetical Project Protocols. This is likely to have enhanced their engagement with, and completion of, the exercise; and should also have strengthened its impact on the successful acquisition of the skills required to design appropriate statistical analyses for causal inference. However it is also plausible that students were more likely to choose contexts and pathways about which they already had strong causal understanding or beliefs, and that these would have made it harder to preference temporality during the subsequent task (covariate classification). This would explain the clinical specialties that were not included in the contexts students chose for their Project Protocol assignments (see Table 1), a number of which only offer placements to medical undergraduates at Leeds Medical School in year 4 or 5 of the MBChB course (Murdoch-Eaton and Roberts, 2009).

### Covariate misclassification prior to DAG specification

Amongst the 71 students who completed the temporality-driven covariate classification task: all (71; 100%) classified at least one covariate as a confounder; fewer (64; 90.1%) classified one or more covariate as a mediator; and fewer still (57; 80.2) classified any covariates as competing exposures. While this is likely to reflect the primary objective of the directed learning exercise – which focused on the identification of potential confounders and likely mediators for obligatory inclusion in (confounders) and exclusion from (mediators) covariate adjustment sets – it is also possible that students felt prompted, or indeed obliged, to specify at least *some* covariates as mediators and competing exposures, even though these are essentially secondary considerations when compiling appropriate covariate adjustment sets containing (only) confounders.

Indeed, through careful assessment of the covariates classified as confounders, mediators and competing exposures it was possible to identify a substantial proportion that had been misclassified (see Table 2). These included 2 covariates that were assessed as equivalent/identical to the specified exposure (one of which had been misclassified as a likely mediator; the other as a competing exposure); and 3 covariates that were assessed as being consequences of the specified outcome (all 3 of which had been misclassified as likely mediators). These errors aside, misclassification rates were lowest (at 4.8%) amongst the 252 covariates classified as potential confounders, although these misclassifications involved a larger proportion of the 71 students who completed this task (at 15.5%). Misclassification was substantially higher amongst the 123 covariates classified as mediators (at 56.9%) and amongst the 137 classified as competing exposures (at 67.9%); and once again these proportions were higher still amongst the tasks completed by the 64 and 57 students who classified one or more of their selected covariates as likely mediators (70.3%) and competing exposures (78.9%), respectively.

**Table 2.** A comparison of student-reported and assessor-validated covariate classifications.

| Assessor-validated covariate classification: | Student-reported covariate classification: |  |  |  |  |  |
| --- | --- | --- | --- | --- | --- | --- |
|  | Confounder<br>(n=71/252) |  | Mediator<br>(n=64/123) |  | Competing exposure<br>(n=57/137) |  |
|  | per student<br>n (%) | per covariate<br>n (%) | per student<br>n (%) | per covariate<br>n (%) | per student<br>n (%) | per covariate<br>n (%) |
| Exposure | 0 (0%) | 0 (0%) | 1 (1.6%) | 1 (0.8%) | 1 (1.8%) | 1 (0.7%) |
| Outcome | 0 (0%) | 0 (0%) | 0 (0%) | 0 (0%) | 0 (0%) | 0 (0%) |
| Confounder | 60 (84.5%) | 241 (95.6%) | 40 (62.5%) | 55 (44.7%) | 37 (64.9%) | 75 (54.7%) |
| Mediator | 11 (15.5%) | 11 (4.4%) | 19 (29.7%) | 53 (43.1%) | 15 (26.3%) | 17 (12.4%) |
| Competing Exposure | 0 (0%) | 0 (0%) | 10 (15.6%) | 11 (8.9%) | 12 (21.1%) | 44 (32.1%) |
| Consequence of the outcome | 0 (0%) | 0 (0%) | 3 (4.7%) | 3 (2.4%) | 0 (0%) | 0 (0%) |

As a result of these errors, only 9 (12.7%) of the students were assessed as having correctly classified all of their selected covariates as confounders, mediators *and/or* competing exposures; although 8 of these students achieved this after classifying their selected covariates *either* as confounders and mediators, *or* as confounders and competing exposures. This left just one student who had correctly classified one or more of their selected covariates as confounder, mediator *and* competing exposure. These findings suggest that while most of the students (71/85; 83.5%) successfully completed the third of the four tasks in the directed learning exercise, very few (9/71; 12.7%) were assessed to have done so correctly; and the vast majority of those that achieved this (8/9; 88/9%) did so without including (and therefore needing to successfully classify) *any* covariates as likely mediators and/or competing exposures. Clearly, the very modest level of success achieved in covariate classification during the temporality-driven covariate classification task (i.e. the third of the directed learning exercise tasks) suggests that the instructions and embedded prompts to privilege temporality over prior causal knowledge/belief were either ineffectual or very difficult for students to apply. Indeed, as mentioned earlier, this may have proved particularly challenging against the backdrop of their existing knowledge, training, beliefs and/or interests in the healthcare contexts and pathways that they themselves had chosen as the basis for their hypothetical Project Protocol assignments.

### Covariate misclassification during DAG specification

Similar types of errors in the misclassification of covariates as potential confounders, likely mediators and competing exposures were also evident in the DAGs dawn by the 66 (77.6%) students who went on to complete the final task of the directed learning exercise. However, it is clear that a substantial proportion of the 58 (68.2%) students who completed both this and the preceding task (temporality-driven covariate classification) altered the classification of their selected covariates when they came to specify their DAGs. Indeed, half (29; 50.0%) of these DAGs contained the same number of covariates classified as potential confounders as those classified as such in the preceding task; while 19 (32.8%) contained more and 10 (17.2%) contained fewer. The equivalent proportions for mediators were: 34 (58.6%) the same; 10 (17.2%) more; and 14 (24.1%) fewer; while for competing exposures: 36 (62.1%) DAGs contained the same number as those specified in the third task; 15 (25.9%) more; and 7 (12.1%) fewer.

While, the total number of covariates classified as likely mediators in the third task (100) was essentially the same as the number included in these 58 DAGs (99); the total number of potential confounders and competing exposures included in the DAGs were around 10% higher than those classified as such in the third task (confounders: 235 vs. 213; competing exposures: 123 vs. 112). These findings indicate that students altered not only the classification of their selected covariates between the third and fourth task of the directed learning exercise, but also the total number of covariates involved. As such, it is clear that DAG specification did not simply involve the placement of covariates within the DAG in line with the classifications made in the preceding task, but that drawing a DAG had prompted students to reconsider *both* the covariates they had chosen (in the second task of the exercise: covariate selection) *and* how these covariates had been classified (in the third task of the exercise: temporality-driven covariate classification). It also seems clear that DAG specification had led the students to re-evaluate the role(s) that (other) potential covariates might play in relation to any potential causal relationship between the exposure and outcome they had chosen in the very first task (exposure and outcome specification). Clearly DAG specification, *in and of itself*, proved to be a task that invoked and involved considerations other than temporality when students decided which covariates were relevant for consideration/inclusion, and what role(s) each of these might play within their DAG.

Further evidence to this effect is available from an assessment of the small number of DAGs (8/66; 12.1%) drawn by students who had completed this task without classifying their covariates in the preceding task (temporality-driven covariate classification). This revealed a far smaller proportion of misclassified covariates (just 3/56 or 5.4%), comprising just one covariate in each of the 3/8 (37.5%) DAGs concerned. All three of these covariates had been misclassified as competing exposures, one of which was assessed as being a likely mediator, and the other two as potential confounders. Unfortunately, a subsample of just 8 DAGs offers scant evidence on which to draw firm conclusions, not least because 3 of these DAGs (all of which were assessed as having no misclassified covariates) had only included covariates classified as potential confounders and either likely mediators (2) or competing exposures (1); and here, as before, it is likely that misclassification rates were lower simply because there were fewer opportunities for error. Nonetheless, the lower proportion of misclassified covariates in this small sample of DAGs is consistent with the view that the thoughts and actions required when specifying a DAG might help to reduce the misclassification of covariates.

Additional support for this proposition can be found in the lower proportion of misclassified covariates amongst the remaining 58 DAGs, each of which had been specified by students who had previously completed the preceding task (temporality-driven covariate classification). Indeed, notwithstanding the inclusion of additional and alternative covariates in these DAGs, the proportion of DAGs with one or more misclassified covariate (37; 63.8%) was substantially lower than that observed from the classification of covariates in the third task of the directed learning exercise (87.3% see Table 2, above), although neither approached the low levels of covariate misclassification achieved by the 8 students who specified their DAGs without completing the third task beforehand.

Further research is therefore warranted to confirm (rather than infer) the reasons and reasoning involved in DAG specification and in the use of DAG specification vs. temporality in covariate classification. Such research will be necessary to establish:

i. the extent to which it is possible to base covariate classification decisions solely on careful and nuanced consideration of the temporal sequence of known covariates that have been conceptualised as characteristics of entities, phenomena and processes, but have been operationalised as either time invariant or time variant measurements; and
ii. whether social conditioning, experiential knowledge and training create such strong cognitive and heuristic prejudices that it can prove impossible to consistently preference temporality over prior causal understanding or belief (Wilson 1983).

Such research might generate important insights simply by conducting in-depth interviews with established analysts who have used temporality-driven covariate classification and/or DAG specification to inform the design of appropriate statistical models that can support causal inference from observational analyses. Experimental psychological techniques might also assist in elucidating the substantial challenges that conceptualisation, operationalisation and cognitive heuristics pose in achieving the phenomenological and critical realist perspectives and insights that are likely to be required to accurately and consistently interpret covariates (and their measurements) as markers of temporally anchored and relational ‘events’ which can then be used to inform the design of statistical analyses capable of informing causal inference.

### DAG specification errors and their likely consequences

Meanwhile, across all of the DAGs specified by the 66 (77.6%) students who completed this final task, the median total number of nodes included was 10 and ranged from 4 to 14 (these numbers comprising all included covariates, *as well as* the specified exposure and specified outcome). All of these DAGs included at least one covariate assessed as representing a potential confounder, and the median number of such nodes was 4 (range: 1-10). In contrast, 17 (25.8%) DAGs contained no likely mediators, and the median number of mediators was just 1 (range: 0-6). Likewise, 9 (13.6%) DAGs contained no competing exposures, and the median number of competing exposures was 2 (range: 0-5).

Interestingly, one student also included two nodes in their DAG that were assessed as being consequences of the outcome, though none of the three students who had previously selected (and misclassified) such covariates during the second and third tasks (see Table 2, above) then went on to represent these as such in their DAGs (i.e. with arcs leading to them from the outcome, with or without additional arcs from the specified exposure and all of the other included covariates). Elsewhere, 16 (24.2%) DAGs used composite ‘super-nodes’ (i.e. a single node with which two or more covariates were associated; Tennant et al. 2020), and in these 16 DAGs the median number of arcs drawn was just 6 (range: 3-9), while for the remaining 50 (75.8%) DAGs – all of which had separate nodes for each of the selected covariates – the median number of arcs was 12 (4-22). Neither approach to DAG specification (using super-nodes or separate nodes) generated DAGs that were assessed as being ‘forward saturated’ (i.e. included all possible arcs between temporally separated nodes); and all but one had missing arcs between covariates specified as potential confounders and those specified as likely mediators. There were also a substantial number of DAGs missing arcs: between confounders and the specified outcome (32; 48.5%); from the specified exposure to any mediators (21; 31.8%); and from any mediators to the specified outcome (23; 34.8%). Indeed, there were even 9 (13.6%) DAGs in which the arc between the specified exposure and the specified outcome was missing.

While a strict interpretation of these missing arcs would have made it challenging to assess which of the DAGs had correctly classified covariates as potential confounders, likely mediators or competing exposures, this was achieved by interpreting: covariates with arcs leading *into* the exposure as confounders; those with arcs leading *out of* the exposure as mediators; and those with only a single arc leading *into* the outcome as competing exposures – an approach that was validated by reference to the classificatory labels which 27 (40.9%) students had included next to individual covariates or clusters of covariates in their DAGs. Thus, despite the fact that almost all of the DAGs contained missing arcs (a serious issue given the strong assumption ‘absent arcs’ imply), only a handful of DAGs contained errors suggesting a fundamental lack of understanding: only one contained a cyclical causal path; only one had used ‘directionless arcs’ (i.e. arcs that lacked arrows); and only one had arcs that ended in the middle of another arc (in this instance the arc between the specified exposure and outcome) rather than ending at one of the nodes at either end of that arc. Yet together with the large number missing arcs and the associated failure to apply or achieve ‘forward saturation’, all of the specified DAGs contained at least one technical error, and none of the students succeeded in applying the DAG specification instructions correctly.

Since the principal utility of specifying DAGs in full (i.e. with every temporally plausible arc included) is to facilitate the identification of covariate adjustment sets for relationships between *any* specified exposure and outcome, the DAGs drawn by students in the present study would have added little value to the classification of covariates undertaken in the preceding task (temporality-driven covariate classification) had this task not incurred extensive misclassification. However, all but a handful (6; 9%) of the specified DAGs provided a sufficiently clear indication of which covariates were considered potential confounders, likely mediators and competing exposures to support the identification of a covariate adjustment set for mitigating the effect of measured confounding when estimating any causal effect between their specified exposure and specified outcome. That said, as was evident from the assessment of the results of the previous task (temporality-driven covariate classification; see Table 2, above), clearly indicating which covariates are considered confounders, mediators or competing exposures offers little benefit to subsequent analytical modelling if these have been incorrectly classified; and although the fourth task (DAG specification) involved lower rates of covariate misclassification to that achieved during the preceding task (temporality-driven covariate classification), misclassification rates were still high enough (at 63.8%) to introduce substantial avoidable bias in any subsequent analyses aiming to estimate the causal relationship between the specified exposure and specified outcome if these had relied upon covariate adjustment sets derived using the covariates as classified in these DAGs.

## Conclusion

The present study extends our understanding of the ease with which directed learning in causal inference techniques might be integrated within undergraduate courses that aim to equip students with practical analytical and statistical skills. Using an exercise designed to provide directed learning in each of the four successive tasks involved in temporality-driven covariate classification and DAG specification, as well as substantial scope for student choice regarding the contexts and topics in which these tasks were applied, the present study demonstrated that high levels of student engagement and task completion can be achieved. However, completion rates declined with each successive task, and a large number of errors were made in the last two tasks (temporality-driven covariate classification; and DAG specification) which would have substantively undermined their analytical utility. Taken together, these findings suggest that some students struggled to complete the exercise in the time available, and that those who completed all four tasks may have only been able to do so at the expense of the diligence required to follow instructions, consolidate their learning and attain a degree of proficiency.

Nonetheless, the additional and alternative covariates selected, together with the improvements in covariate classification, that occurred as a result of completing the final task (DAG specification) confirms the important contribution that drawing causal path diagrams can make in elucidating the covariate adjustment sets required to mitigate bias from measured confounders in analyses of observational data that aim to support causal inference. This was an unexpected finding, not least because temporality is arguably the only objective basis upon which probabilistic causal relationships can be determined or defined in observational studies. Indeed, temporality-driven covariate classification (the third task) had been deliberately introduced into the directed learning exercise to address the subjectivity (and lack of consensus) that can arise from reliance on prior knowledge, experience or beliefs regarding causal and functional relationships (Ellison et al. 2014b).

Of course it is possible that the apparent *improvements* in covariate classification observed following DAG specification in the present study simply reflected the tougher conceptual challenge involved in the application of temporality during the preceding task (temporality-driven covariate classification), particularly since the undergraduates involved had limited expertise in the conceptualisation and operationalisation of quantitative variables and the impact thereof on the opacity of temporal relationships amongst and between these. Indeed, these are also exacting challenges for competent analysts with advanced training and substantial experience (Tennant et al. 2020); and there are well-established (if contentious and contested) concerns that drawing DAGs might actually obfuscate rather than elucidate the critical insights and associated thinking required to design analytical models capable of supporting causal inference with observational data (e.g. Krieger and Davey Smith 2016). Clearly, further research is warranted to strengthen our understanding of whether, and how, training in DAG specification (and related techniques) might actually improve the selection of appropriate covariate adjustment sets for use in the analysis of observational data to support causal inference.

## Data Availability

The data analysed in this report are available upon request from the author.

## Acknowledgements

This study would not have been possible without the participation of the MBChB students involved, and the support of colleagues within Leeds School of Medicine, including the Co-Lead of the RESS3 module (Mark Iles), and the module’s Tutorial Supervisors, each of whom make a substantial contribution to the annual programme of analytical training for third year MBChB students. Likewise, the development of undergraduate training in the use of temporality and directed acyclic graphs to develop the skills required to improve the design of statistical models for causal inference has benefitted enormously from collaboration with Johannes Textor (from Radboud University Medical Centre in The Netherlands), and with colleagues from Leeds Causal Inference Group including: Kellyn Arnold, Laurie Berrie, Mark Gilthorpe, Wendy Harrison, John Mbotwa, Peter Tennant and Carol Wilson – all of whom have been forthright in their views, and unstintingly generous with their insights, instincts and ideas.

